# LLM-generated patient-friendly discharge letters for newly diagnosed rheumatic and musculoskeletal diseases: a prospective feasibility study

**DOI:** 10.64898/2026.11.23.26368739

**Authors:** Phillip Kremer, Jana Klemm, Pia Wallmeier, Marie-Therese Holzer, Jessica Pauz, Tingting Xiong, Ina Kötter, Simon Melderis, Isabell Haase, Martin Krusche

## Abstract

**Background:** A new diagnosis of a rheumatic and musculoskeletal disease (RMD) creates substantial information needs, with medical discharge letters often serving as a primary source of information for patients. This study aimed to assess the feasibility, content quality and patient-reported usefulness of large language model (LLM)-generated patient-friendly discharge letters in rheumatology inpatient care.

**Methods:** In this prospective, single-centre, single-arm feasibility study, standard discharge letters were transformed into patient-friendly versions using GPT models, reviewed by treating physicians and provided alongside the originals. We assessed physician review time, corrections, information fidelity, readability, understandability and patient-reported evaluations at follow-up.

**Results:** Of 58 enrolled patients, 52 (89.7%) completed follow-up. Compared with the original letters, patient-friendly letters showed significantly higher Flesch Reading Ease scores (mean difference 11.4, 95% confidence interval 9.8 to 12.9) and higher understandability scores on the Patient Education Materials Assessment Tool (+10.0 percentage points, 95% confidence interval 6.8 to 13.4). Physicians rated all 58 unedited drafts as “very good” or “good”, and review took less than five minutes for 43/58 (74.1%) letters. Unedited LLM drafts contained eleven medical discrepancies in 10/58 (17.2%) letters, ten of which were corrected before discharge. In the final physician-reviewed letters, 310/311 (99.7%) applicable letter–domain assessments were classified as correctly retained or simplified. At follow-up, 42/51 (82.4%) patients rated perceived comparative comprehensibility favourably and 44/51 (86.3%) wished to receive similar letters in future.

**Conclusions:** Physician-reviewed, LLM-generated discharge letters were feasible to implement in routine rheumatology care, with limited physician review time and high patient acceptance. Although text-based readability and assessed understandability improved, objective patient comprehension and clinical benefit remain to be established. Discrepancies in unedited drafts underscore the need for physician oversight.

**Trial registration:** German Clinical Trials Register (DRKS), DRKS00036980. Registered 26 May 2025.

## Background

Patients newly diagnosed with a rheumatic or musculoskeletal disease (RMD) face substantial informational challenges as they must understand their condition, navigate new treatment regimens and assume responsibility for disease self-management [1]. Providing clear and accessible health information is therefore essential, particularly given the high prevalence of limited health literacy (HL) among adults and its association with poorer health outcomes [2–5]. Accordingly, patient education is recommended as an integral part of RMD management, supporting disease knowledge, self-efficacy, self-management and informed participation in shared decision-making [1, 6, 7].

Hospital discharge represents a critical transition in care, particularly for patients newly diagnosed with an RMD. Discharge letters provide essential information on diagnoses, treatment and follow-up care, yet their complexity often exceeds patients’ reading and comprehension abilities [8]. Medical terminology and abbreviations further limit patients’ ability to understand and act on relevant information [7–9]. Poor understanding of discharge information is associated with adverse events and readmissions [11]. These challenges are particularly relevant in rheumatology, where patients need to understand, for example, complex glucocorticoid tapering schedules, infection risks associated with immunosuppressive therapy, laboratory monitoring and warning signs requiring prompt medical attention [4, 6]. Ensuring that discharge information is both comprehensible and medically accurate is therefore important to support safe and effective self-management.

Large language models (LLMs) offer a potentially scalable approach to transforming complex clinical information into patient-friendly language. In rheumatology, LLMs have demonstrated potential in various applications including diagnostic decision support, patient-oriented information and communication [12–15]. They can also simplify complex clinical documents and improve readability and perceived understandability, although factual inaccuracies, omissions and potentially misleading simplifications remain important safety concerns [16, 17]. Previous evaluations of LLM-generated patient-friendly discharge summaries have only been conducted outside rheumatology, for instance in cardiology and internal medicine [16–18].

Patients newly diagnosed with an RMD represent a particularly relevant population for evaluating this approach, given their substantial informational needs, potentially complex treatment regimens and the importance of effective self-management after hospital discharge. Therefore, we conducted to our knowledge the first prospective feasibility study of the integration of LLM-generated patient-friendly discharge letters into routine clinical practice, focussing on workflow feasibility, clinical content quality, patient-reported usefulness and adherence-relevant information needs after discharge.

## Methods

### Study design and setting

This single-centre, prospective, single-arm feasibility study was conducted at the University Medical Center Hamburg-Eppendorf. Recruitment ran from May 2025 to August 2026 and follow-up from August 2025 to October 2026. Where applicable, reporting followed the CONSORT extension for pilot and feasibility trials [19] and the TRIPOD-LLM guideline [20] (Supplementary Materials 1 and 2). The study was registered in the German Clinical Trials Register (DRKS00036980; registration date: 26 May 2025).

### Participants

Consecutive inpatients aged ≥18 years with sufficient German language skills and an RMD first diagnosed during the index admission were eligible. Exclusion criteria were cognitive impairment, a fulminant disease course requiring short-term follow-up and no planned follow-up at our centre. Physician-reported feasibility outcomes at discharge (letter generation, review time, corrections and medical discrepancies) were analysed for all 58 enrolled patients, whereas text-based analyses (readability, PEMAT-P understandability and information fidelity) and patient-reported outcomes were restricted to the 52 patients who completed the follow-up assessment (T1).

### Intervention

Routine discharge letters were manually standardised and processed via UHHGPT [21], an institutional interface to OpenAI models. Input data were not used for model training. Owing to interface updates, three models were used successively (GPT-4o mini, GPT-4.1 nano and GPT-5.4 nano; access periods and number of letters per model in ***Supplementary Table S1***). Two standardised zero-shot prompts (Supplementary Material 3) were applied separately to the medical history and the discharge summary without re-prompting. The outputs were combined with a standard institutional header. The treating physician checked this draft against the original letter and corrected and documented any errors.

### Data collection and outcomes

Data were collected at discharge (T0) and at outpatient follow-up approximately 8–12 weeks later (T1), using study-specific physician and patient questionnaires (Supplementary Material 4). Feasibility was described using letter generation and delivery, correction requirements, recorded physician review time, patient acceptance and follow-up retention.

Primary outcomes comprised physician-rated content quality and comparative comprehensibility at T0, and patient-rated comparative comprehensibility and perceived usefulness at T1. Content quality was rated by the treating physician as an overall judgement of the initial, unedited LLM draft on the German school grading scale (1, very good, to 6, insufficient; lower values indicating better quality). Comparative comprehensibility relative to the original letter was assessed using identically worded five-category items for physicians and patients, with response options ranging from “not at all” to “very well”. Ratings were coded from 1 to 5, with higher values indicating more favourable assessments. Secondary outcomes included readability, PEMAT-P understandability, information fidelity, physician corrections, generation and review time, patient-reported disease understanding and eHealth literacy. The latter was assessed using the validated German eight-item eHealth Literacy Scale (eHEALS; range 8–40), analysed continuously and categorised descriptively as low (8–20), moderate (21–30) or high (31–40) [22–24].

### Outcome assessment

For readability and understandability, the physician-approved patient letter was compared with the medical history and discharge summary sections of the original letter. Readability was assessed using the Amstad adaptation of the Flesch Reading Ease (FRE-Amstad; higher = easier) and the first Vienna formula for non-fiction texts (Erste Wiener Sachtextformel, WSTF; lower = easier) [25, 26]. Understandability was assessed with the Patient Education Materials Assessment Tool for printable materials (PEMAT-P; 17 understandability items [1–12, 15–19]), scored as the percentage of applicable items rated ‘agree’ (0–100%; higher is better) [27]. Two raters (PK, JK) scored all letters independently and resolved disagreements by consensus (prior item-level agreement: unweighted Cohen’s κ = 0.42).

From the medical history and discharge summary of each original letter, we extracted clinically relevant information units (e.g. drug dosage) in eight predefined domains (Supplementary Table S2). Each unit in the physician-approved LLM-generated letter was classified as correctly retained, correctly simplified, omitted or altered (Supplementary Table S3). Unsupported additions (statements absent from the original letter) were recorded separately. Information preservation in the physician-approved letters was summarised at letter–domain level as the proportion of applicable domain assessments classified as correctly retained or simplified. A domain was classified as correctly retained or simplified only if all information units within it were reproduced completely and accurately. A single omitted or altered unit rendered the entire domain discrepant, irrespective of the remaining units. This domain-level measure does not represent the proportion of individual information units preserved or the accuracy of unedited LLM output. Omissions, alterations and unsupported additions were additionally cross-checked against the documented physician corrections.

Documented physician corrections were assigned to one of six predefined categories, ranging from linguistic or stylistic changes to unsupported additions (***Supplementary Table S4***). Safety relevance was assessed independently of correction type as a separate severity criterion: a correction was classified as a safety-relevant error if the uncorrected statement or omission could plausibly have led to patient harm if acted upon, for example incorrect medication or dosing information. We report corrections per letter and the proportion of letters with ≥1 correction.

### Statistical analysis

Analyses were primarily descriptive. Paired comparisons used the Wilcoxon signed-rank test, with mean differences and 95% bootstrap confidence intervals. Patient–physician agreement was assessed using quadratic weighted Cohen’s κ. Exploratory associations between readability changes, patient characteristics and patient-reported outcomes were examined using Spearman’s ρ, with Holm adjustment for multiple testing. Given the exploratory feasibility design, no formal hypothesis-based sample size calculation was performed. The sample size was determined pragmatically by the expected number of eligible patients during the predefined recruitment period. Missing data were not imputed; analyses used available cases with outcome-specific denominators. Analyses were performed using R (version 4.5.2; R Foundation for Statistical Computing, Vienna, Austria) in RStudio (version 2025.09.2+418). Pseudonymised data were stored on secure hospital servers.

The study was conducted in accordance with the Declaration of Helsinki and approved by the local ethics committee in Hamburg, Germany (2025-101483-BO-ff), and all participants provided written informed consent.

During manuscript preparation, the authors used ChatGPT (GPT-5.5; OpenAI) to improve the readability and language of the text. The authors reviewed and edited all output and take full responsibility for the content of the publication.

## Results

Of 58 enrolled patients with a newly diagnosed RMD, six (10.3%) did not complete the follow-up assessment. Physician-reported feasibility outcomes at discharge are reported for all 58 patients, and text-based and patient-reported analyses for the 52 patients (89.7%) who completed T1 (***Figure 1***). The median age was 67.5 years (IQR 54.3–75.8), and 30 (51.7%) were female. The most frequent diagnoses were large-vessel vasculitis (n=21, 36.2%), rheumatoid arthritis (n=11, 19.0%) and connective tissue disease (n=7, 12.1%). Patient characteristics, educational attainment and self-reported digital health literacy of all 58 enrolled patients are summarised in ***Table 1***; the corresponding data for the 52 patients who completed follow-up are provided in ***Supplementary Table S5***.

**Figure 1:**
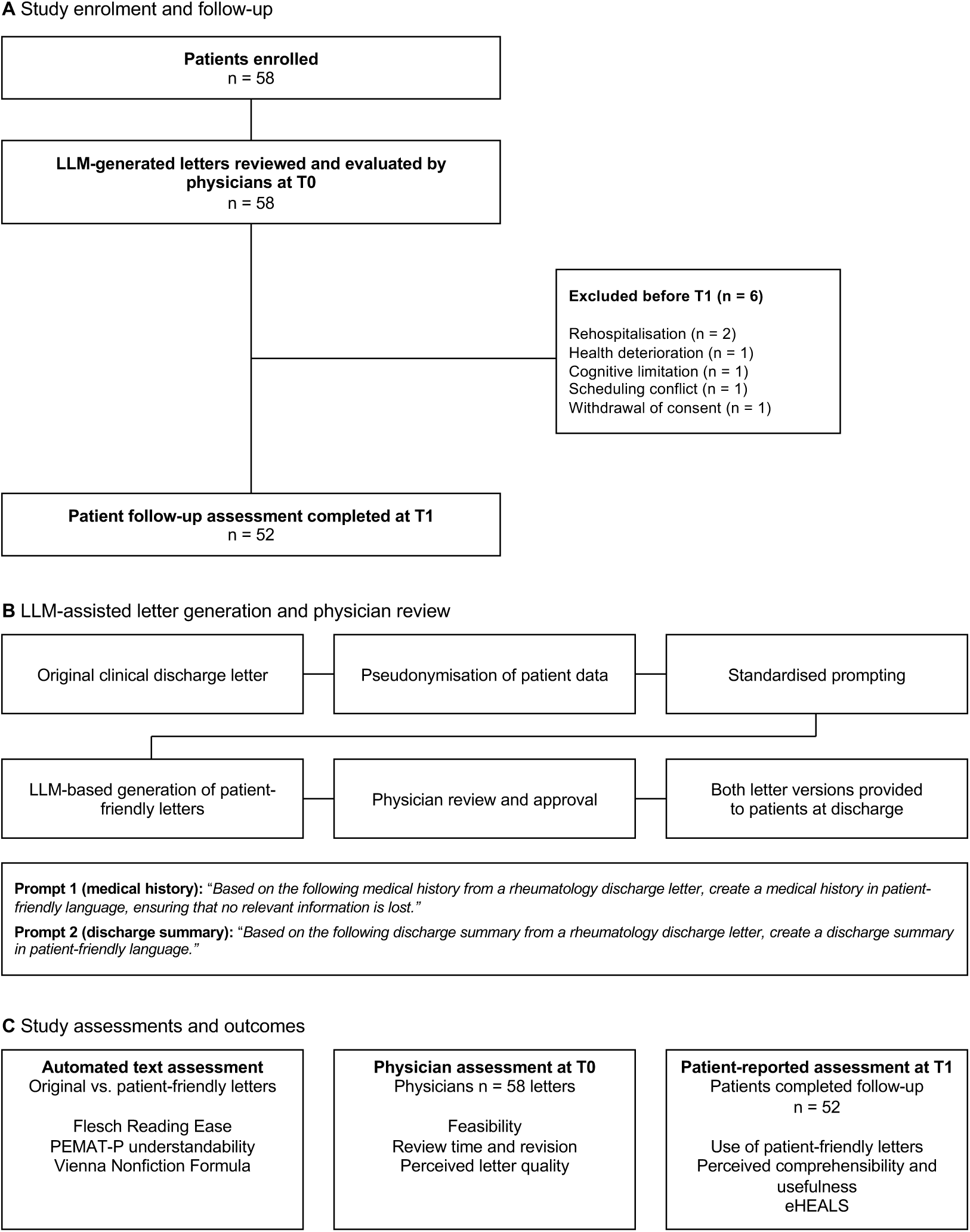
Study flow, letter generation workflow and evaluation. **(a)** Patient flow: of 58 enrolled patients, six were excluded before T1 and 52 were included in text-based and patient-reported analyses. **(b)** Generation of patient-friendly letters from pseudonymised discharge letters using standardised prompts, followed by physician review and approval. **(c)** Outcome assessment by automated text analysis, physician survey at discharge (T0) and patient survey at follow-up (T1)

**Table 1:** Patient characteristics of all enrolled patients (n = 58)

| Characteristic | Group | Total n (%) |
|---|---|---|
| **Gender (n = 58)** | Female | 30 (51.7) |
|  | Male | 28 (48.3) |
| **Age,** years **(n = 58)** | 18-35 | 8 (13.8) |
|  | 36-45 | 4 (6.9) |
|  | 46-55 | 4 (6.9) |
|  | 56-65 | 11 (19.0) |
|  | 66-75 | 16 (27.6) |
|  | >75 | 15 (25.9) |
|  | Median (IQR) | 67.5 (54.3-75.8) |
| **Disease (n = 58)** | Large vessel vasculitis | 21 (36.2) |
|  | Rheumatoid arthritis | 11 (19.0) |
|  | Connective tissue disease | 7 (12.1) |
|  | ANCA-associated vasculitis | 5 (8.6) |
|  | Polymyalgia rheumatica | 5 (8.6) |
|  | Psoriatic arthritis | 2 (3.4) |
|  | Sarcoidosis | 2 (3.4) |
|  | Still's disease | 2 (3.4) |
|  | Gout | 1 (1.7) |
|  | IgG4-related disease | 1 (1.7) |
|  | Reactive arthritis | 1 (1.7) |
| **Education (n = 51)** | No education qualification | 1 (2.0) |
|  | Lower education qualification | 11 (21.6) |
|  | Intermediate education qualification | 18 (35.3) |
|  | Higher education qualification | 13 (25.5) |
|  | University degree | 8 (15.7) |
| **Health literacy,**<br>*eHEALS score* **(n = 52)** | Low | 11 (21.2) |
|  | Moderate | 24 (46.2) |
|  | High | 17 (32.7) |

Notes. Values are n (%) of all enrolled patients (N = 58) unless otherwise stated. Educational attainment was available for 51 patients. eHEALS was assessed at follow-up (T1) and is therefore available only for the 52 patients who completed the follow-up assessment; percentages refer to these denominators. eHEALS categories: low 8–20, moderate 21–30, high 31–40. Abbreviations: eHEALS, eHealth Literacy Scale; IQR, interquartile range.

Per patient, the original discharge letters listed a median of five (IQR 3–6) newly initiated medications and four (IQR 3–5) recommendations for further disease management (e.g. vaccination). Physician review-time data were available for 58/58 (100%) letters and eHEALS data for 52/58 (89.7%), and 51/58 (87.9%) patients completed the letter-related follow-up questionnaire, at a median of 10.1 weeks (IQR 8.7–12.3).

Compared with the original letters, the physician-approved patient-friendly versions showed significantly improved readability and understandability across linguistic measures (***Figure 2***). Mean Flesch Reading Ease scores increased from 30.8 (SD 5.20) to 42.1 (SD 5.27), with a mean paired difference of 11.36 points (95% bootstrap CI 9.77 to 12.88; p<0.001). Mean First Vienna Non-Fiction Text Formula scores decreased from 13.5 (SD 0.94) to 12.0 (SD 0.94), corresponding to a difference of −1.54 (95% bootstrap CI −1.82 to −1.24; p<0.001). Mean PEMAT understandability scores increased from 43.0% to 53.1%, with a paired improvement of 10.01 percentage points (95% bootstrap CI 6.80 to 13.35; p<0.001). The most frequently improved PEMAT criteria were use of common, everyday language (fulfilled in 0 vs 29 letters), division of information into short sections (7 vs 25) and clear presentation of numbers (23 vs 38).

**Figure 2:**
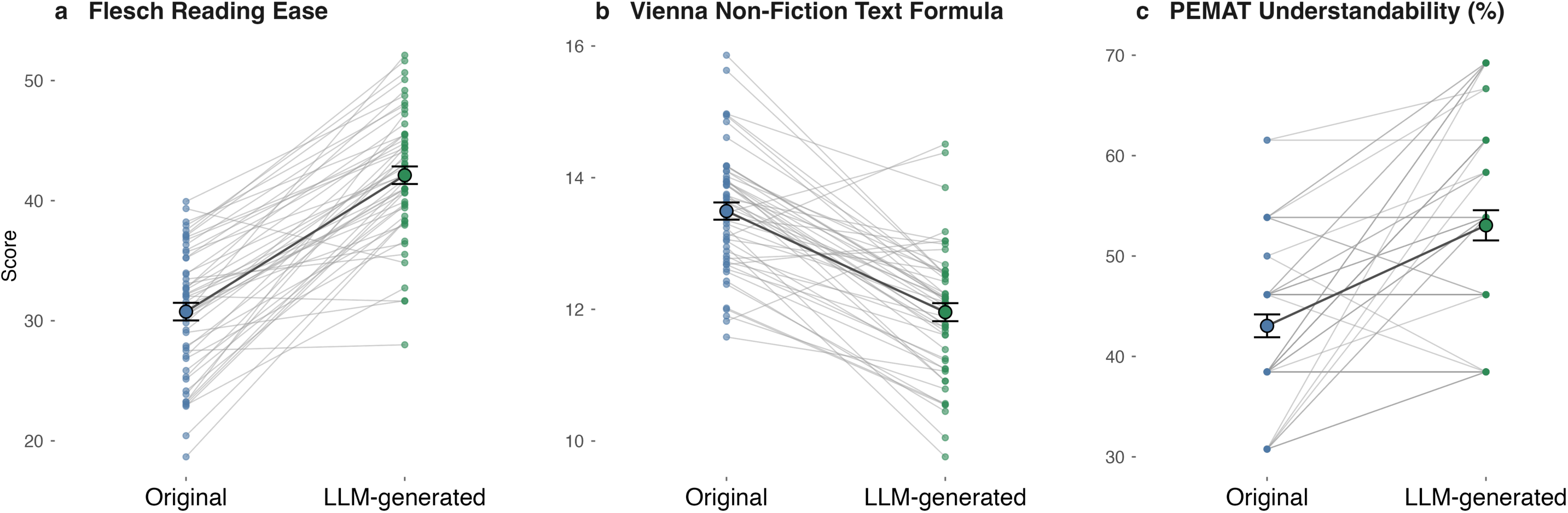
Readability and understandability of original and patient-friendly letters. **(a)** Flesch Reading Ease (higher = easier). **(b)** First Vienna Non-Fiction Text Formula (lower = easier). **(c)** PEMAT-P understandability for original and physician-approved LLM-generated letters (n=52).

Treating physicians rated the content quality of all 58 initial, unedited LLM drafts as either “very good” (grade 1, 43/58 [74.1%]) or “good” (grade 2, 15/58 [25.9%]). Generation of the patient-friendly letter required <5 min for 10/58 (17.2%) letters, 5–10 min for 47/58 (81.0%) and 20–30 min for 1/58 (1.7%); none required 10–20 min. Physician review required <5 min for 43/58 (74.1%) letters and 5–10 min for 15/58 (25.9%); none required >10 min (***Figure 3a***). Overall, 24/58 (41.4%) letters required no correction and 34/58 (58.6%) required at least one (Figure 3b), with a total of 61 corrections. Of these 61 corrections, 37 (60.7%) were linguistic or stylistic and 14 (23.0%) were clarifications of otherwise correct content. Together, 51/61 (83.6%) concerned language or clarity. In contrast, 10/61 (16.4%) addressed medical content (none meeting the criterion for a safety-relevant error), comprising four omissions, five factual inaccuracies and one unsupported addition (***Supplementary Table S4***).

**Figure 3.**
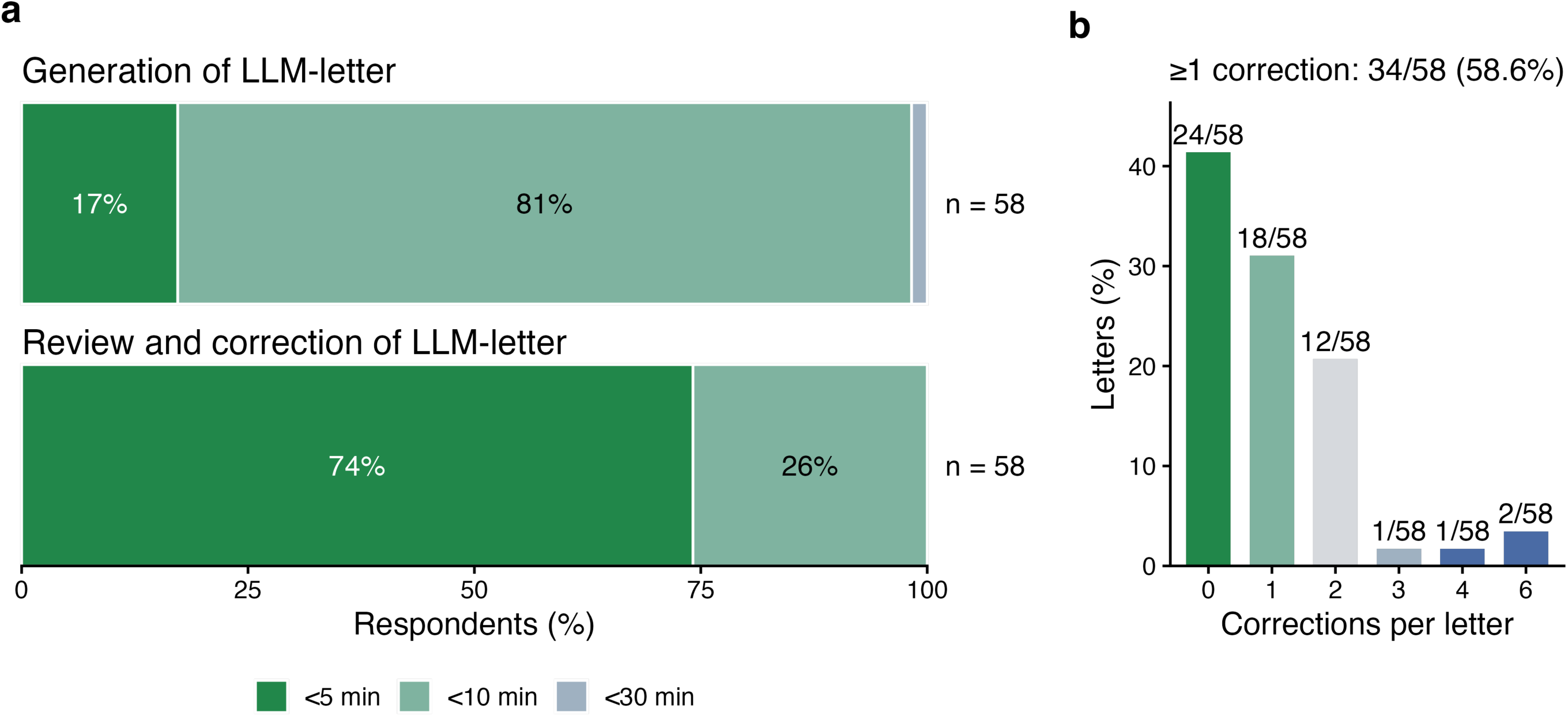
Time requirements and editing burden of LLM-assisted discharge letter generation. **(a)** Physician-reported time required to generate the patient-friendly letter and to review it before approval (n=58 letters). **(b)** Proportion of letters requiring no correction or at least one correction during physician review (n=58).

Information fidelity was evaluated across eight clinical domains in the edited LLM output after physician review. After excluding non-applicable domains, 311 letter–domain assessments remained. Of these, 248 (79.7%) were correctly retained and 62 (19.9%) were correctly simplified, yielding a domain-level preservation rate of 99.7% in the physician-reviewed letters. Simplifications occurred most frequently in the new diagnosis and new medication domains (***Figure 4***).

**Figure 4:**
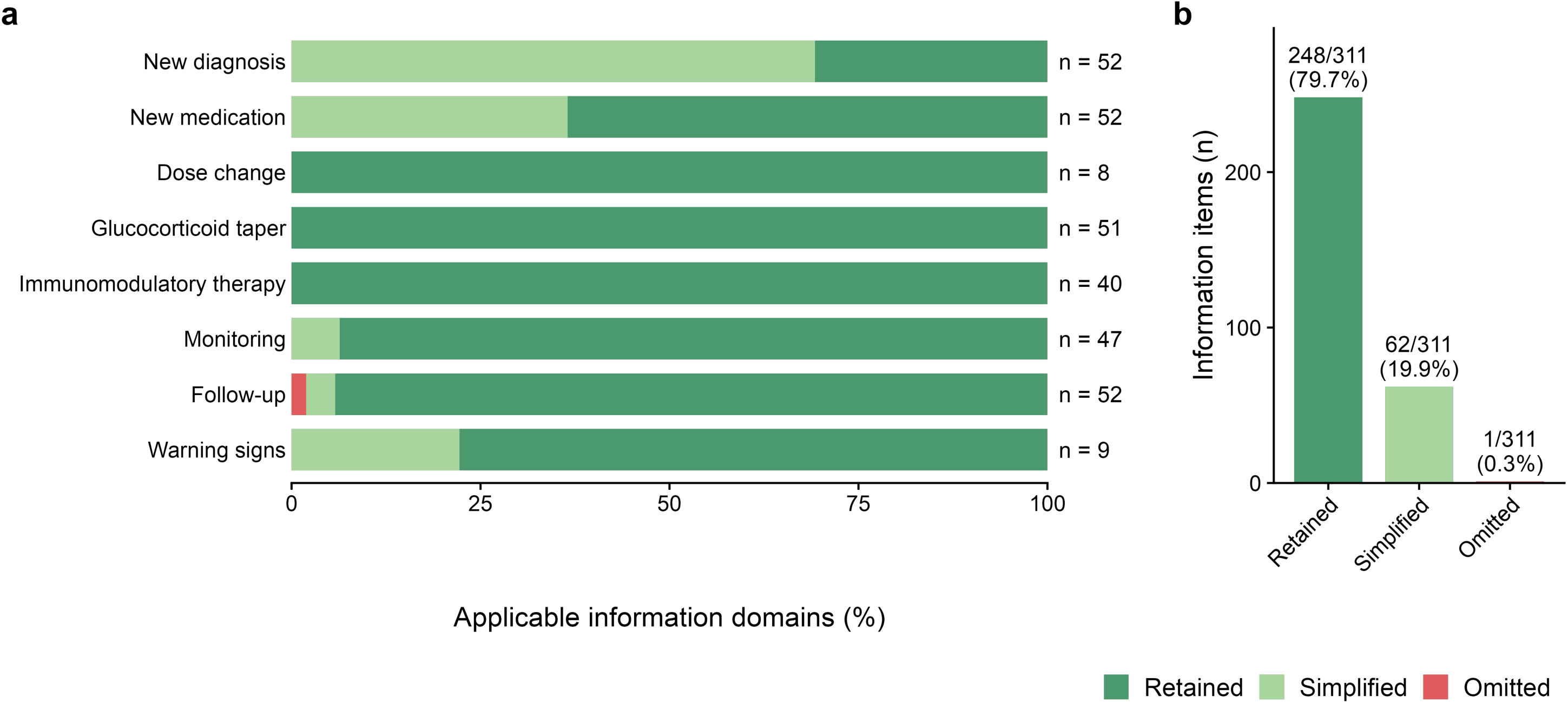
Preservation of clinical information in physician-reviewed LLM-generated discharge letters. **(a)** Proportion of letter–domain assessments per clinical domain classified as correctly retained, correctly simplified, omitted or altered in the physician-approved LLM-generated letters; n indicates the number of letters in which the domain was applicable. **(b)** Overall distribution of all letter–domain assessments across categories (n=311 assessments from 52 letters across eight predefined domains; non-applicable domains excluded).

In the unedited LLM drafts, physicians identified ten medical discrepancies in 9/58 (15.5%) letters before discharge (***Supplementary Table S4***), e.g. a missing referral to the outpatient clinic or an added symptom. One further omission (a planned gastroscopy) was missed during review and identified only after discharge; it was not classified as safety-relevant and is not included in Supplementary Table S4. Overall, the unedited LLM output thus contained 11 discrepancies in 10/58 (17.2%) letters. These affected the domains new diagnosis (6/11), follow-up (4/11) and new medication (1/11).

At T1, 51/51 (100.0%) respondents reported reading their patient-friendly letter. The letter was rated as helpful for understanding the diagnosis by 36/51 (70.6%) and for understanding the next steps in care by 41/51 (80.4%). After discharge, 26/51 (51.0%) patients searched for further information on the content of their letter, most frequently on the internet (24/51, 47.1%) and from their general practitioner (14/51, 27.5%). Overall, 44/51 (86.3%) expressed a preference for receiving a patient-friendly version alongside the standard discharge letter in the future. Response distributions and item-specific denominators are shown in Figure 5.

**Figure 5:**
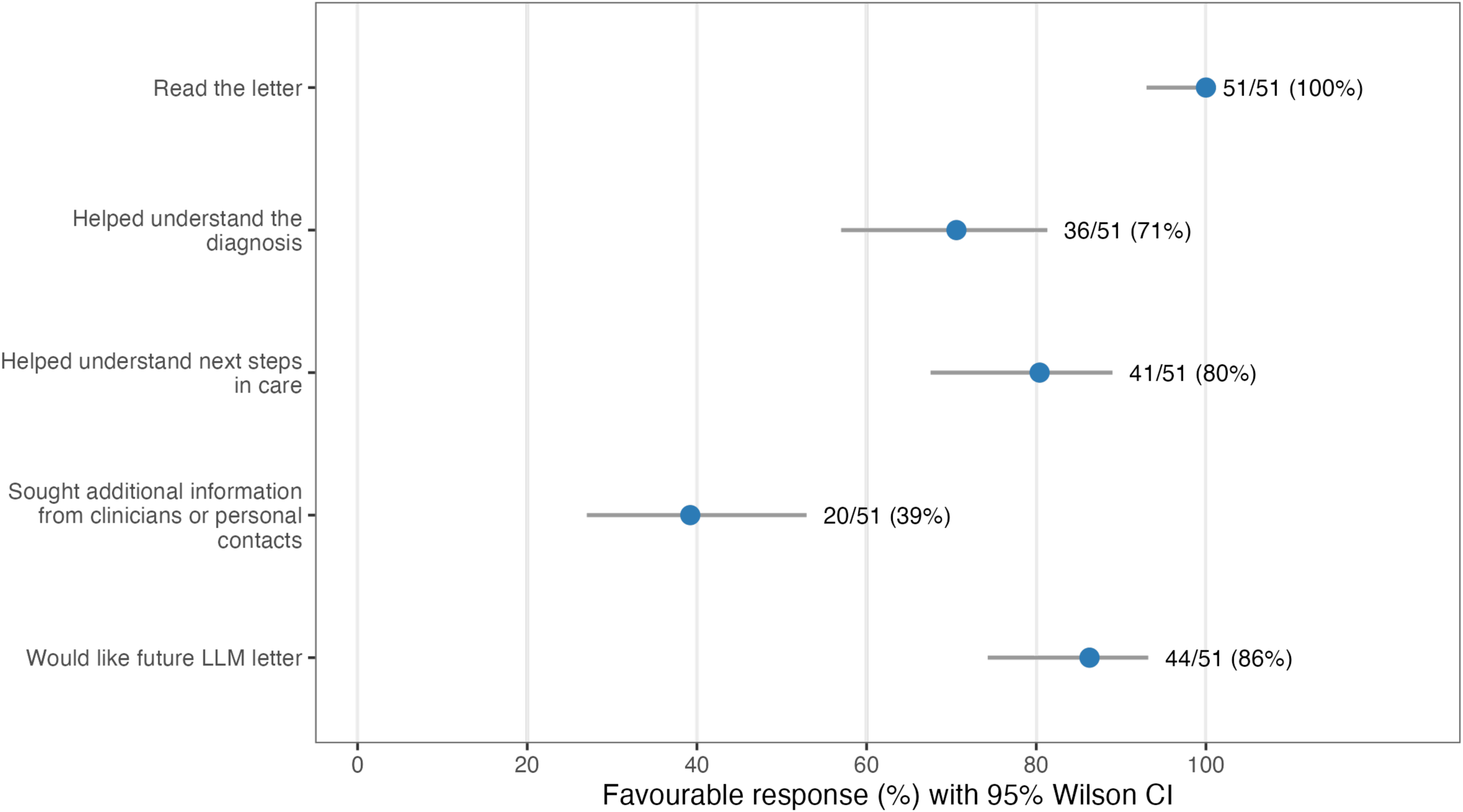
Patient-reported use and perceived value at T1. Proportions of patients endorsing each statement at follow-up, with 95% Wilson confidence intervals. Patient-reported evaluations were available for 51 of 52 participants; one patient did not provide responses to the letter evaluation items.

Treatment-related information needs persisted after discharge. Before the outpatient follow-up visit (T1), 19/51 (37.3%) patients contacted a physician, usually their general practitioner (16/19) and rarely a specialist (2/19) or the ward physician (2/19; multiple responses possible), most frequently because of questions regarding side effects (n = 4), medication (n = 3), glucocorticoid tapering (n = 2) and vaccination status (n = 2). In total, 12 of 23 free-text reasons concerned newly initiated treatment (side effects, medication, glucocorticoid tapering, vaccination or additional medication) (***Supplementary Table S6***). Overall, 44/51 (86.3%) expressed a preference for receiving a patient-friendly version alongside the standard discharge letter in the future. Response distributions and item-specific denominators are shown in ***Figure 5***.

Patients and treating physicians rated the comparative comprehensibility of the patient-friendly letters on a five-point scale at follow-up and discharge, respectively (***Figure 6***). At follow-up, 42/51 (82.4%) patients rated the comprehensibility of the patient-friendly letter relative to the original letter as “well” or “very well”. Patients rated the letters less favourably than physicians in 31/51 cases (60.8%), equally in 15/51 (29.4%), and more favourably in 5/51 (9.8%). Nevertheless, ratings differed by no more than one scale point in 44/51 cases (86.3%). The mean paired difference (patient minus physician) was −0.71 points (Wilcoxon signed-rank test, p < 0.001). Quadratic weighted agreement was low (κ = 0.06, 95% CI −0.13 to 0.21).

**Figure 6:**
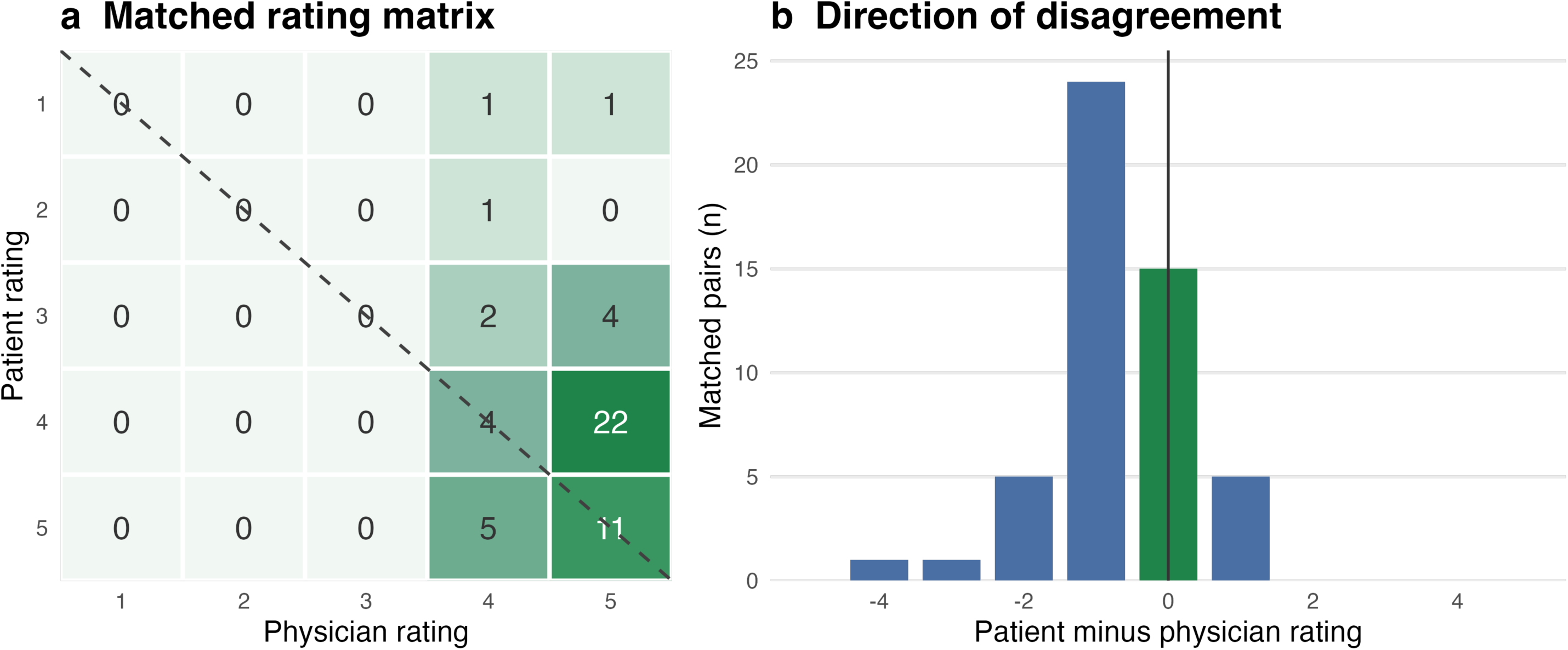
Differences in perceived comprehensibility between patients and physicians. **(a)** Paired ratings of the perceived comparative comprehensibility of physician-reviewed, LLM-generated patient-friendly discharge letters relative to standard discharge letters. Physicians provided their assessments at discharge (T0), whereas patients were surveyed at follow-up (T1). Cell values indicate the number of patient–physician rating pairs, and the diagonal represents identical ratings. **(b)** Direction of differences between paired patient and physician ratings.

In exploratory analyses, changes in FRE, WSTF and PEMAT scores between original and patient-friendly letters were not significantly associated with patient-reported understandability or usefulness at follow-up (n = 51; Spearman’s ρ −0.24 to 0.16; all Holm-adjusted p ≥ 0.518). Confidence intervals included zero for all six correlations (Supplementary Table S7). Regarding patient characteristics, higher self-reported eHealth literacy was associated with higher patient-rated comprehensibility (n = 51; Spearman’s ρ = 0.40; Holm-adjusted p = 0.035). Higher educational attainment was associated with lower perceived usefulness of the LLM-generated letters (n = 50; Spearman’s ρ = −0.38; Holm-adjusted p = 0.0498). Education was not significantly associated with perceived understandability (n = 50; Spearman’s ρ = 0.22; Holm-adjusted p = 0.73). All remaining correlations, including all those involving age, were weak and non-significant (all |ρ| ≤ 0.25; all Holm-adjusted p ≥ 0.59; Supplementary Table S8).

## Discussion

To our knowledge, this is the first prospective study evaluating physician-reviewed, LLM-generated patient-friendly discharge letters in routine rheumatology inpatient care. The workflow required limited recorded physician review time, patient-reported acceptance was high, and approved letters demonstrated improved readability and PEMAT-P understandability. These findings are relevant to newly diagnosed RMD patients, who frequently face complex treatment regimens and substantial information needs. In our cohort, this need was pronounced: patients were predominantly older (median age 67.5 years), and two thirds had complex systemic diseases (vasculitis, connective tissue disease, Still’s disease, sarcoidosis or IgG4-related disease; 38/58, 65.5%). At discharge, patients had been prescribed a median of five newly initiated medications and four further care recommendations.

First, LLM-assisted letter generation was feasible within the investigated workflow, with physician review requiring less than five minutes for 43/58 (74.1%) letters, and 24/58 (41.4%) letters requiring no corrections. However, 34/58 (58.6%) required corrections, including ten involving medical content. In the unedited LLM-generated drafts, eleven medical discrepancies were identified in 10/58 letters (17.2%). Ten were corrected during physician review, while one omission was identified retrospectively after discharge. Following physician review, 310 of 311 applicable clinical domain assessments (99.7%) were classified as correctly retained or simplified. This domain-level measure reflects the final physician-reviewed documents and should not be interpreted as the error rate of the unedited LLM output. Simplifications were most frequent in the new diagnosis and new medication domains, which are particularly relevant to newly diagnosed patients.

Previous studies have similarly identified omissions, inaccuracies and unsupported additions in LLM-generated discharge information [16, 28, 29] including medication-related safety issues in older and more complex patients [30]. Our findings extend this evidence by demonstrating that clinical discrepancies can occur despite otherwise favourable readability and workflow characteristics. Although none of our detected discrepancies met the predefined criterion for safety relevance, one omission remained undetected until after discharge. Our small study therefore cannot establish the safety of either unreviewed or physician-reviewed output. Four of eleven discrepancies concerned follow-up and six diagnosis, underscoring the need to verify actionable information.

Second, the patient evaluations were predominantly favourable: 44/51 (86.3%) patients indicated that they would welcome similar letters in the future, while 36/51 (70.6%) and 41/51 (80.4%) found them helpful for understanding their diagnosis and next steps in care, respectively. At follow-up, 42 of 51 patients (82.4%) rated the comprehensibility of the patient-friendly letters relative to the original discharge letters as “well” or “very well”. These observations are consistent with a recent randomised trial reporting improved patient activation following physician-reviewed LLM-generated discharge summaries [18]. However, patient preference and perceived comprehensibility should not be interpreted as evidence of improved objective disease understanding.

Importantly, patients and treating physicians evaluated the comparative comprehensibility of the patient-friendly letters favourably. Although patients rated the letters less favourably than physicians in 31/51 (60.8%) paired assessments, 44/51 (86.3%) ratings differed by at most one scale point. Agreement was low (κ = 0.06), suggesting that physician ratings may not adequately capture patients’ perspectives. This supports involving patients directly in the evaluation of patient-facing LLM output, as in our previous patient-derived benchmark in connective tissue diseases [31]. As physicians rated letters at discharge and patients at follow-up, differences may reflect timing and recall.

Nevertheless, further information needs persisted as about half of the patients (26/51, 51.0%) sought additional information and 19/51 (37.3%) contacted a physician before the follow-up visit. Their questions mainly concerned side effects, laboratory results, medication, glucocorticoid tapering and vaccinations, which are practical aspects of newly initiated treatment rather than the diagnosis itself. Given the number of new medications and recommendations per patient, these findings identify persistent information needs, but do not establish how completely the letters addressed these needs or whether they changed subsequent information-seeking behaviour. Future versions could address the above-mentioned treatment-related topics more explicitly. Moreover, they could direct patients to reliable online resources, given the frequent use of the internet.

Third, readability and structural understandability improved compared with standard letters (mean FRE difference +11.36 points, PEMAT-P +10.01 percentage points). These results are consistent with previous studies reporting improvements in the readability of LLM-simplified clinical documents [16, 17, 32, 33]. However, the observed improvements reflect the whole workflow, not the LLM alone. Moreover, readability formulas and PEMAT scores assess characteristics of written information rather than patients’ actual comprehension. In our exploratory analyses, improvements in these readability or understandability measures were not significantly associated with patient-reported comprehensibility or usefulness. Given the small sample size and imprecise correlation estimates, this does not exclude an association but highlights the need for objective comprehension measures.

Exploratory analyses suggested that higher self-reported eHealth literacy was associated with more favourable comprehensibility ratings, whereas lower educational attainment was associated with greater perceived usefulness (Holm-adjusted p = 0.035 and p = 0.0498, respectively). These distinct subjective outcomes cannot establish differential objective benefit. In particular, eHEALS assesses self-perceived digital health literacy rather than general health literacy or comprehension of printed information [22, 34]. These small-sample findings are hypothesis-generating and warrant investigation in larger, more diverse patient populations.

Study limitations include the small single-centre sample size, six losses to follow-up and the single-arm, unblinded design, which precludes causal inference. Study-specific patient questionnaires completed weeks after discharge introduce recall and measurement bias, and physicians assessed letters they had reviewed themselves. We did not measure objective comprehension, adherence or clinical outcomes. Model changes during recruitment and continuing LLM development may further limit transferability.

## Conclusions

In conclusion, physician-reviewed, LLM-generated patient-friendly discharge letters were feasible to implement in routine rheumatology inpatient care and were perceived as understandable and useful by patients with a newly diagnosed RMD. By providing accessible and individualised information on diagnoses, treatment and follow-up care, this approach represents a promising strategy to complement conventional discharge communication and support patient education. It may thereby facilitate patient engagement and informed self-management, including adherence to newly initiated treatment, particularly during the transition from hospital to outpatient care. Controlled multicentre studies should evaluate objective understanding, validated adherence measures and clinical outcomes and the effectiveness of structured physician quality assurance.

## Data Availability

All data produced in the present study are available upon reasonable request to the authors.

## Abbreviations

CI: confidence interval
CONSORT: Consolidated Standards of Reporting Trials
DRKS: German Clinical Trials Register (Deutsches Register Klinischer Studien)
eHEALS: eHealth Literacy Scale
FRE: Flesch Reading Ease
GPT: generative pre-trained transformer
HL: health literacy
IQR: interquartile range
LLM: large language model
PEMAT-P: Patient Education Materials Assessment Tool for printable materials
RMD: rheumatic and musculoskeletal disease
SD: standard deviation
T0: discharge assessment
T1: follow-up assessment
TRIPOD-LLM: Transparent Reporting of a multivariable prediction model for Individual Prognosis Or Diagnosis – Large Language Models
WSTF: first Vienna formula for non-fiction texts (Erste Wiener Sachtextformel).

## Declarations

### Ethics approval and consent to participate

The study was conducted in accordance with the Declaration of Helsinki and approved by the local ethics committee in Hamburg, Germany (2025-101483-BO-ff). All participants provided written informed consent.

### Consent for publication

Not applicable.

### Availability of data and materials

The datasets generated and analysed during the current study are not publicly available owing to the sensitive nature of the clinical information and applicable data-protection restrictions, but are available from the corresponding author on reasonable request. Aggregated results supporting the findings are presented in the manuscript and its supplementary information files.

### Authors’ contributions

Conception and design of the study: PK, IH, MK. Recruitment of patients and acquisition of data: PK, JK, PW, MTH, JP, TX, SM. Generation and physician review of patient-friendly discharge letters: PK, PW, MTH. Statistical analysis: JK, PK. Interpretation of data: PK, JK, MK. Drafting of the manuscript: PK, JK, MK. Supervision: IK, IH, MK. All authors critically reviewed and approved the final manuscript and agree to be accountable for all aspects of the work. PK is the guarantor of the study and accepts full responsibility for the work and had access to the data.

## Acknowledgements

Not applicable.

