## Supplementary for "LLM-generated patient-friendly discharge letters for newly diagnosed rheumatic and musculoskeletal diseases: a prospective feasibility study"

### Large language model-generated patient-friendly discharge letters for newly diagnosed rheumatic and musculoskeletal diseases: a prospective feasibility study - Supplementary Material

#### Material 1. CONSORT checklist for pilot and feasibility trials

##### Reporting according to the CONSORT 2010 extension to randomised pilot and feasibility trials, adapted for a non-randomised, single-arm pilot and feasibility study

| Item | Reporting item | Page |
|---|---|---|
| **Title and abstract** | | |
| 1a | Title identifies the study as a pilot or feasibility trial | 1 |
| 1b | Structured summary of design, methods, results and conclusions | 2 |
| **Introduction** | | |
| 2a | Scientific background; rationale for a future definitive trial and for the pilot | 4, 11 |
| 2b | Specific objectives or research questions of the pilot | 4 |
| **Methods** | | |
| 3a | Description of pilot study design | 5 |
| 3b | Important changes to methods after commencement, with reasons | 5 (model changes) |
| 4a | Eligibility criteria for participants | 5 |
| 4b | Settings and locations of data collection | 5 |
| 4c | How participants were identified and consented | 5, 7 |
| 5 | Interventions in sufficient detail to allow replication, incl. how and when administered | 5; Suppl. M3 |
| 6a | Prespecified assessments addressing each pilot objective, incl. how and when assessed | 5–7 |
| 6b | Changes to assessments after commencement, with reasons | NR |
| 6c | Prespecified progression criteria for a future definitive trial, if applicable | NR |
| 7a | Rationale for the sample size number | 7 |
| 7b | Interim analyses and stopping guidelines, if applicable | NR |
| 8a | Generation of the random allocation sequence | N/A |
| 8b | Type of randomisation and any restriction | N/A |
| 9 | Allocation concealment mechanism | N/A |
| 10 | Who generated the sequence, who enrolled and assigned participants | 5, 12 |
| 11a | Blinding, if done: who was blinded and how | 11 |
| 11b | Similarity of interventions, if relevant | N/A |
| 12 | Methods used to address each pilot objective (qualitative or quantitative) | 6–7 |
| **Results** | | |
| 13a | Participant flow: approached/assessed, enrolled, received the intervention, assessed per objective | 7–9, 16 |

| Item | Reporting item | Page |
|---|---|---|
| 13b | Losses and exclusions after enrollment, with reasons | 7, 16 |
| 14a | Dates of recruitment and follow-up | 5 |
| 14b | Why the pilot ended or was stopped | 7 |
| 15 | Baseline demographic and clinical characteristics | 7, 15 |
| 16 | Number of participants included in each analysis (per objective) | 7–9, 15–19 |
| 17 | Results per objective, with measures of uncertainty (e.g., 95% CI) | 7–9, 17–19 |
| 18 | Other analyses that may inform the future definitive trial | 9 |
| 19 | Important harms or unintended effects | 8, 9–10 |
| 19a | Other important unintended consequences, if relevant | NR |
| **Discussion** | | |
| 20 | Limitations, sources of bias and remaining uncertainty about feasibility | 11 |
| 21 | Generalisability of methods and findings to a future trial and other studies | 10–11 |
| 22 | Interpretation consistent with objectives and findings, balancing benefits and harms | 9–11 |
| 22a | Implications for progression to a definitive trial, incl. proposed amendments | 11 |
| **Other information** | | |
| 23 | Trial registration number and name of trial registry | 5 |
| 24 | Where the protocol can be accessed, if available | NR |
| 25 | Funding sources and role of funders | 12 |
| 26 | Ethical approval with reference number | 7 |

Notes. Abbreviated CONSORT items adapted from Eldridge et al. (BMJ. 2016;355:i5239). Page numbers refer to the 20-page V10 manuscript; NR = not reported; N/A = not applicable; Suppl. = supplementary material.

#### Material 2. TRIPOD-LLM checklist

##### Reporting according to the TRIPOD-LLM guideline for studies using large language models

| Item | Reporting item | Research design | LLM task | Page |
|---|---|---|---|---|
| **Title and abstract** | | | | |
| 1 | Title identifies development/tuning/evaluation of an LLM, the task, population and outcome | All | All | 1 |
| 2 | Abstract per TRIPOD-LLM for Abstracts | All | All | 2 |
| **Introduction** | | | | |
| 3a | Healthcare context/use case and rationale, with reference to existing approaches | All | All | 4, 9–11 |

| Item | Reporting item | Research design | LLM task | Page |
|---|---|---|---|---|
| 3b | Target population, intended use in the care pathway and intended users | E, H | All | 4–5 |
| 4 | Study objectives, incl. development, fine-tuning and/or validation stage of LLM | All | All | 4 |
| **Methods** | | | | |
| 5a | Data sources for training, tuning and/or evaluation, with rationale | All | All | 5 |
| 5b | Relevant data points and their distribution; dataset descriptors (e.g., language, country) | All | All | 5, 7, 15 |
| 5c | Dates of oldest and newest text used for development and evaluation datasets | All | All | 5 (study period) |
| 5d | Data pre-processing and quality checks, incl. consistency across sources and groups | All | All | 5–6, 16 |
| 5e | Handling of missing and imbalanced data; reasons for omitting data | All | All | 5, 7 |
| 6a | LLM name, version and last date of training | All | All | 5; Suppl. S1 |
| 6b | LLM development: architecture, training, fine-tuning, alignment strategy and goals | M, D | All | N/A |
| 6c | Text generation: prompt engineering, output consistency, inference settings | M, D, E | All | 5; Suppl. M3 |
| 6d | Initial and post-processed LLM output | All | All | 5–6, 8 |
| 6e | Classification details, probability derivation and thresholds | All | C, OF | N/A |
| 7a | Metrics for quality of generative output and errors compared with a reference standard | All | QA, IR, DG, SS, MT | 6, 8 |
| 7b | Relevance of outcome metrics to the downstream task; correlation with human evaluation | E, H | All | 6–7, 9 |
| 7c | Outcome definition, how predictions were obtained, inference date (closed-source), metrics | E, H | All | 5–6, 8–9 |
| 7d | Subjective assessment: assessor qualifications, instructions, demographics, agreement | All | All | 5–6, 9; Suppl. M4 |
| 7e | Comparison with other LLMs, humans or benchmarks | All | All | 6–9 |
| 8a | Annotation process and guidelines, with examples | All | All | 6; Suppl. S2–S4 |
| 8b | Number of annotators, proportion double-annotated, inter-annotator agreement | All | All | 6 |
| 8c | Background/experience of annotators or characteristics of labelling models | All | All | 6, 12 |

| Item | Reporting item | Research design | LLM task | Page |
|---|---|---|---|---|
| 9a | Prompt design, curation and selection process | All | All | 5; Suppl. M3 |
| 9b | Data used to develop the prompts | All | All | NR |
| 10 | Pre-processing of data before summarisation | All | SS | 5, 16 |
| 11 | Instruction tuning/alignment: instructions, data, interface, evaluator characteristics | M, D | All | N/A |
| 12 | Compute requirements or proxies (time, cost, hardware, inference time) | M, D, E | All | 8, 17 |
| 13 | Ethics committee approval and informed consent or waiver | All | All | 7 |
| **Open science** | | | | |
| 14a | Funding sources and role of funders | All | All | 12 |
| 14b | Conflicts of interest and financial disclosures | All | All | 12 |
| 14c | Access to the study protocol, or statement that none was prepared | H | All | NR |
| 14d | Registration details, or statement that the study was not registered | H | All | 5 |
| 14e | Availability of study data | All | All | NR |
| 14f | Availability of code to reproduce results | All | All | NR |
| **Patient and public involvement** | | | | |
| 15 | Patient and public involvement, or statement of no involvement | H | All | NR |
| **Results** | | | | |
| 16a | Flow of documents/participants through the study, incl. follow-up | E, H | All | 7, 16 |
| 16b | Characteristics overall and per data source/setting, incl. key dates and sample size | E, H | All | 5, 7, 15; Suppl. S5 |
| 16c | Comparison of clinical variables between development and evaluation data, if available | E, H | All | N/A |
| 16d | Number of participants and outcome events in each analysis | E, H | All | 7–9, 17–19 |
| 17 | LLM performance on prespecified metrics (7a) and/or human evaluation (7d) | All | All | 7–9, 17–19 |
| 18 | Results of any LLM updating, if applicable | All | All | N/A |
| **Discussion** | | | | |
| 19a | Overall interpretation, incl. fairness, in the context of objectives and prior work | All | All | 9–11 |
| 19b | Limitations and their effect on bias, uncertainty and generalisability | All | All | 11 |

| Item | Reporting item | Research design | LLM task | Page |
|---|---|---|---|---|
| 19c | Known challenges with data for the task and domain (representation, missingness, bias) | E, H | All | 10–11 |
| 19d | Intended use: input, end-user and level of autonomy/human oversight | E, H | All | 5–6, 10–11 |
| 19e | Handling of poor-quality or unavailable input data in clinical use, if applicable | E, H | All | NR |
| 19f | Required user interaction and expertise, if applicable | E, H | All | 5–6, 9–11 |
| 19g | Next steps for future research, regarding applicability and generalisability | All | All | 11 |

Notes. Abbreviated TRIPOD-LLM items from Gallifant et al. (Nat Med. 2025;31:60–69). Page numbers refer to the 20-page V10 manuscript; NR = not reported; N/A = not applicable; Suppl. = supplementary material.

#### Material 3. LLM-generation prompt

**Prompt 1 – German: „***Erstelle mir basierend auf der folgenden Anamnese aus einem rheumatologischen Arztbrief eine Anamnese in Patienten-freundlicher Sprache ohne das relevante Informationen verloren gehen:“*

**Prompt 1 – English:** *“Based on the following medical history from a rheumatology discharge letter, create a medical history in patient-friendly language, ensuring that no relevant information is lost.”*

**Prompt 2 – German: “***Erstelle mir basierend auf der folgenden Epikrise aus einem rheumatologischen Arztbrief eine Epikrise in Patienten-freundlicher Sprache:“*

**Prompt 2 – English:** *“Based on the following discharge summary from a rheumatology discharge letter, create a discharge summary in patient-friendly language.”*

#### Material 4. Questionnaires

##### Physician questionnaire at T0 in German:

1. Wie lange haben Sie für das Verfassen des ursprünglichen Entlassberichtes benötigt?

☐ < 15 min ☐ 15–30 min ☐ 30–60 min ☐ 1–2 h ☐ > 2 h

2. Wie lange haben Sie für die Generierung des UHHGPT Arztbriefes benötigt?

☐ < 5 min ☐ < 10 min ☐ < 20 min ☐ < 30 min ☐ > 30 min

3. Wie lange haben Sie nach der Generierung benötigt, um den UHHGPT Arztbrief Korrektur zu lesen und ggf. zu korrigieren?

☐ < 5 min ☐ < 10 min ☐ < 20 min ☐ < 30 min ☐ > 30 min

4. Falls Sie Korrekturen durchführen mussten: Welche waren dies (z. B. grammatikalisch, fachliche Aspekte, neu generierte Informationen etc.)? Bitte benennen Sie bei fachlichen Aspekten die konkrete Korrektur, z.B. falsche Dosierung, andere Empfehlungen etc.

5. Wie viel mussten Sie prozentual gesehen vom UHHGPT Arztbrief noch korrigieren?

☐ < 5 % ☐ < 10 % ☐ < 20 % ☐ < 30 % ☐ < 50 % ☐ > 50 %

6. Wie gut fanden Sie den initial von UHHGPT entworfenen Arztbrief im Schulnotensystem (1: sehr gut, 6: ungenügend)?

☐ 1 ☐ 2 ☐ 3 ☐ 4 ☐ 5 ☐ 6

7. Im Vergleich zum ursprünglichen Entlassbericht: Fanden Sie den UHHGPT Arztbrief besser verständlich?

☐ sehr gut ☐ gut ☐ mittel ☐ wenig ☐ gar nicht

8. Sehen Sie in der Implementierung des UHHGPT generierten Entlassungsbriefes einen zusätzlichen Nutzen in der klinischen Praxis (1: stimme vollkommen zu, 6: stimme gar nicht zu)?

☐ 1 ☐ 2 ☐ 3 ☐ 4 ☐ 5 ☐ 6

#### Physician questionnaire at T0 in English:

1. How long did it take you to write the original discharge letter?

☐ < 15 min ☐ 15–30 min ☐ 30–60 min ☐ 1–2 h ☐ > 2 h

2. How long did it take you to generate the UHHGPT letter?

☐ < 5 min ☐ < 10 min ☐ < 20 min ☐ < 30 min ☐ > 30 min

3. After generation, how long did it take you to proofread and, if necessary, correct the UHHGPT letter?

☐ < 5 min ☐ < 10 min ☐ < 20 min ☐ < 30 min ☐ > 30 min

4. If you had to make corrections, what were they (e.g. grammatical, medical content, newly generated information)? For medical content, please specify the exact correction, e.g. incorrect dosage, different recommendations.

5. What percentage of the UHHGPT letter did you still have to correct?

☐ < 5 % ☐ < 10 % ☐ < 20 % ☐ < 30 % ☐ < 50 % ☐ > 50 %

6. How would you rate the letter initially drafted by UHHGPT on the German school grading scale (1: very good, 6: insufficient)?

☐ 1 ☐ 2 ☐ 3 ☐ 4 ☐ 5 ☐ 6

7. Compared with the original discharge letter: did you find the UHHGPT letter easier to understand?

☐ very well ☐ well ☐ moderately ☐ a little ☐ not at all

8. Do you see an additional benefit for clinical practice in implementing the UHHGPT-generated discharge letter (1: strongly agree, 6: strongly disagree)?

☐ 1 ☐ 2 ☐ 3 ☐ 4 ☐ 5 ☐ 6

#### Patient questionnaire at T1 in German:

Geschlecht:

☐ männlich ☐ weiblich ☐ divers

Alter: ____________

Höchster Bildungsabschluss:

☐ ohne Schulabschluss
☐ Hauptschulabschluss
☐ Realschulabschluss/Mittlere Reife
☐ Fachhochschulreife/Abitur
☐ Hochschulabschluss

1. Wie nützlich ist das Internet Ihrer Meinung nach, um Ihnen bei Entscheidungen über Ihre Gesundheit zu helfen?

☐ sehr nützlich ☐ nützlich ☐ unsicher ☐ wenig nützlich ☐ überhaupt nicht nützlich

2. Wie wichtig ist es für Sie, auf Gesundheitsressourcen im Internet zugreifen zu können?

☐ sehr wichtig ☐ wichtig ☐ unsicher ☐ wenig wichtig ☐ überhaupt nicht wichtig

3. Ich weiß, welche Gesundheitsressourcen im Internet verfügbar sind.*

☐ stimme voll zu ☐ stimme zu ☐ unsicher ☐ stimme nicht zu ☐ stimme überhaupt nicht zu

4. Ich weiß, wo ich hilfreiche Gesundheitsressourcen im Internet finden kann.*

☐ stimme voll zu ☐ stimme zu ☐ unsicher ☐ stimme nicht zu ☐ stimme überhaupt nicht zu

5. Ich weiß, wie ich hilfreiche Gesundheitsressourcen im Internet finden kann.*

☐ stimme voll zu ☐ stimme zu ☐ unsicher ☐ stimme nicht zu ☐ stimme überhaupt nicht zu

6. Ich weiß, wie ich das Internet nutzen kann, um meine Fragen zur Gesundheit zu beantworten.*

☐ stimme voll zu ☐ stimme zu ☐ unsicher ☐ stimme nicht zu ☐ stimme überhaupt nicht zu

7. Ich weiß, wie ich die Gesundheitsinformationen, die ich im Internet finde, für mich nutzen kann.*

☐ stimme voll zu ☐ stimme zu ☐ unsicher ☐ stimme nicht zu ☐ stimme überhaupt nicht zu

8. Ich habe die Fähigkeiten, die ich brauche, um die im Internet gefundenen Gesundheitsressourcen zu bewerten.*

☐ stimme voll zu ☐ stimme zu ☐ unsicher ☐ stimme nicht zu ☐ stimme überhaupt nicht zu

9. Ich kann hochwertige Gesundheitsressourcen von minderwertigen Gesundheitsressourcen im Internet unterscheiden.*

☐ stimme voll zu ☐ stimme zu ☐ unsicher ☐ stimme nicht zu ☐ stimme überhaupt nicht zu

10. Ich fühle mich sicher bei der Nutzung von Informationen aus dem Internet, um gesundheitliche Entscheidungen zu treffen.*

☐ stimme voll zu ☐ stimme zu ☐ unsicher ☐ stimme nicht zu ☐ stimme überhaupt nicht zu

*Die folgenden Fragen beziehen sich auf den Entlassungsbericht/Arztbrief, den Sie am Entlassungstag erhalten haben:*

11. Haben Sie Ihren stationären Arztbrief gelesen?

☐ ja ☐ nein

12. Wie gut haben Sie den Arztbrief verstanden?

☐ sehr gut ☐ gut ☐ mittel ☐ wenig ☐ gar nicht

13. Haben Sie nach weiteren Informationen zu den Inhalten des Arztbriefes gesucht?

☐ ja ☐ nein

14. Falls ja, wo und wie lange haben Sie jeweils danach gesucht? (Mehrfachantworten möglich)

| | **< 1 h** | **1–2 h** | **3–5 h** | **5–10 h** | **> 10 h** |
|---|---|---|---|---|---|
| Internet | ☐ | ☐ | ☐ | ☐ | ☐ |
| Fachbücher | ☐ | ☐ | ☐ | ☐ | ☐ |
| Andere Fachärzt:innen | ☐ | ☐ | ☐ | ☐ | ☐ |
| Hausärzt:in | ☐ | ☐ | ☐ | ☐ | ☐ |
| Bekanntenkreis | ☐ | ☐ | ☐ | ☐ | ☐ |
| Andere medizinische Fachberufe (z. B. Apotheker:innen, Physiotherapeut:innen) | ☐ | ☐ | ☐ | ☐ | ☐ |

15. Wie gut haben Sie sich durch den Arztbrief informiert gefühlt und wussten, was Sie bis zu Ihrem Ambulanztermin tun sollten (z. B. Medikamenteneinnahme, weitere Termine/Untersuchungen etc.)?

☐ sehr gut ☐ gut ☐ mittel ☐ wenig ☐ gar nicht

16. Mussten Sie vor Ihrem Ambulanztermin nochmal Kontakt zu Ärzt:innen aufnehmen, da Sie Nachfragen hatten?

☐ ja ☐ nein

17. Falls ja, zu welchen?

☐ Hausärzt:in ☐ Fachärzt:in ☐ Stationsärzt:in

18. Falls ja, weshalb?

19. Wie gut haben Sie sich durch Ihren Arztbrief auf den ersten Ambulanztermin vorbereitet gefühlt?

☐ sehr gut ☐ gut ☐ mittel ☐ wenig ☐ gar nicht

20. Haben Sie Inhalte aus dem Arztbrief während des Ambulanztermins besprochen (z. B. Laborwerte, Fachbegriffe)?

☐ ja ☐ nein

21. Falls ja, welche?

*Die folgenden Fragen beziehen sich auf den durch UHHGPT formulierten Arztbrief, den Sie per Post zugeschickt bekommen haben:*

22. Haben Sie den durch UHHGPT formulierten Arztbrief gelesen?

☐ ja ☐ nein

23. Falls ja, wie gut haben Sie ihn verstanden?

☐ sehr gut ☐ gut ☐ mittel ☐ wenig ☐ gar nicht

24. Im Vergleich zum ursprünglichen Entlassbericht: Fanden Sie den UHHGPT Arztbrief besser verständlich?

☐ sehr gut ☐ gut ☐ mittel ☐ wenig ☐ gar nicht

25. Haben Sie das Gefühl, dass Sie durch den UHHGPT Arztbrief umfassender und besser über Ihre Erkrankung informiert sind?

☐ sehr gut ☐ gut ☐ mittel ☐ wenig ☐ gar nicht

26. Würden Sie es als hilfreich empfinden, wenn Sie in Zukunft bei Arztbesuchen zusätzlich einen durch UHHGPT formulierten Arztbrief erhalten würden?

☐ sehr gut ☐ gut ☐ mittel ☐ wenig ☐ gar nicht

** Items der eHealth Literacy Scale (eHEALS; doi: 10.2196/jmir.8.4.e27).*

#### Patient questionnaire at T1 in English:

Sex:

☐ male ☐ female ☐ diverse

Age: ____________

Highest educational qualification:

☐ no school-leaving qualification
☐ lower secondary school certificate (Hauptschulabschluss)
☐ intermediate secondary school certificate (Realschulabschluss/Mittlere Reife)
☐ higher education entrance qualification (Fachhochschulreife/Abitur)
☐ university degree

1. How useful do you feel the Internet is in helping you make decisions about your health?

☐ very useful ☐ useful ☐ unsure ☐ not very useful ☐ not useful at all

2. How important is it for you to be able to access health resources on the Internet?

☐ very important ☐ important ☐ unsure ☐ not very important ☐ not important at all

3. I know what health resources are available on the Internet.*

☐ strongly agree ☐ agree ☐ undecided ☐ disagree ☐ strongly disagree

4. I know where to find helpful health resources on the Internet.*

☐ strongly agree ☐ agree ☐ undecided ☐ disagree ☐ strongly disagree

5. I know how to find helpful health resources on the Internet.*

☐ strongly agree ☐ agree ☐ undecided ☐ disagree ☐ strongly disagree

6. I know how to use the Internet to answer my questions about health.*

☐ strongly agree ☐ agree ☐ undecided ☐ disagree ☐ strongly disagree

7. I know how to use the health information I find on the Internet to help me.*

☐ strongly agree ☐ agree ☐ undecided ☐ disagree ☐ strongly disagree

8. I have the skills I need to evaluate the health resources I find on the Internet.*

☐ strongly agree ☐ agree ☐ undecided ☐ disagree ☐ strongly disagree

9. I can tell high-quality health resources from low-quality health resources on the Internet.*

☐ strongly agree ☐ agree ☐ undecided ☐ disagree ☐ strongly disagree

10. I feel confident in using information from the Internet to make health decisions.*

☐ strongly agree ☐ agree ☐ undecided ☐ disagree ☐ strongly disagree

*The following questions refer to the discharge letter you received on the day of discharge:*

11. Did you read your inpatient discharge letter?

☐ yes ☐ no

12. How well did you understand the discharge letter?

☐ very well ☐ well ☐ moderately ☐ a little ☐ not at all

13. Did you look for further information about the contents of the discharge letter?

☐ yes ☐ no

14. If yes, where did you look and for how long in each case? (multiple answers possible)

| | **< 1 h** | **1–2 h** | **3–5 h** | **5–10 h** | **> 10 h** |
|---|---|---|---|---|---|
| Internet | ☐ | ☐ | ☐ | ☐ | ☐ |
| Medical reference books | ☐ | ☐ | ☐ | ☐ | ☐ |
| Other specialist physicians | ☐ | ☐ | ☐ | ☐ | ☐ |
| General practitioner | ☐ | ☐ | ☐ | ☐ | ☐ |
| Friends and acquaintances | ☐ | ☐ | ☐ | ☐ | ☐ |
| Other healthcare professionals (e.g. pharmacists, physiotherapists) | ☐ | ☐ | ☐ | ☐ | ☐ |

15. How well informed did you feel by the discharge letter, and did you know what to do until your outpatient appointment (e.g. taking medication, further appointments/examinations)?

☐ very well ☐ well ☐ moderately ☐ a little ☐ not at all

16. Did you have to contact a physician again before your outpatient appointment because you had questions?

☐ yes ☐ no

17. If yes, which physician?

☐ general practitioner ☐ specialist physician ☐ ward physician

18. If yes, why?

19. How well prepared for your first outpatient appointment did you feel as a result of your discharge letter?

☐ very well ☐ well ☐ moderately ☐ a little ☐ not at all

20. Did you discuss contents of the discharge letter during the outpatient appointment (e.g. laboratory values, medical terms)?

☐ yes ☐ no

21. If yes, which?

*The following questions refer to the letter written by UHHGPT that was sent to you by post:*

22. Did you read the discharge letter written by UHHGPT?

☐ yes ☐ no

23. If yes, how well did you understand it?

☐ very well ☐ well ☐ moderately ☐ a little ☐ not at all

24. Compared with the original discharge letter: did you find the UHHGPT letter easier to understand?

☐ very well ☐ well ☐ moderately ☐ a little ☐ not at all

25. Do you feel that the UHHGPT letter has informed you more comprehensively and better about your condition?

☐ very well ☐ well ☐ moderately ☐ a little ☐ not at all

26. Would you find it helpful to receive an additional letter written by UHHGPT after future medical appointments?

☐ very well ☐ well ☐ moderately ☐ a little ☐ not at all

** Items of the eHealth Literacy Scale (eHEALS; doi: 10.2196/jmir.8.4.e27).*

#### Supplementary Table S1. Distribution of LLM-generated plain-language discharge letters by model in the analysis cohort

| Model | Model-use period | Letters, n (%) |
|---|---|---|
| GPT-4o mini | Study start through 9 December 2025 | 26 (50.0%) |
| GPT-4.1 nano | 10 December 2025 through 7 May 2026 | 10 (19.2%) |
| GPT-5.4 nano | 8 May 2026 through 14 August 2026 | 16 (30.8%) |

#### Supplementary Table S2. Clinical information load of the original letter
##### Variables extracted per patient from the final original discharge letter

| Variable | Operationalization |
|---|---|
| New diagnoses | Number of newly established main and relevant secondary diagnoses during the index stay |
| New medications | Number of new substances started at discharge due to the new RMD or its management |
| Dose changes | Number of relevant dose changes to existing medications |
| Glucocorticoid taper | Yes/no; additionally, number of described taper steps |
| Immunomodulatory therapy | Yes/no; e.g., csDMARD, b/tsDMARD, cyclophosphamide, rituximab |
| Monitoring | Laboratory, blood pressure, vaccination, infection or other safety checks |
| Follow-up | Outpatient clinic appointments, general practitioner appointments, specialist check-ups, imaging, etc |
| Warning signs | Explicitly stated red flags or situations requiring earlier medical presentation |

*Abbreviations*: RMD, rheumatic and musculoskeletal disease; csDMARD, conventional synthetic disease-modifying antirheumatic drug; b/tsDMARD, biological/targeted synthetic disease-modifying antirheumatic drug.

#### Supplementary Table S3. Coding scheme for the content fidelity of LLM-generated plain-language discharge letters compared with the original letters

| Category | Code | Definition |
|---|---|---|
| Correctly retained | retained | Content is present in full, clinically accurate and with identical meaning |
| Correctly simplified | simplified | Content rephrased in lay language without loss of meaning |
| Omitted | omitted | Information missing entirely |
| Altered | altered | Information was changed |
| Added | added | LLM adds a clinical statement that cannot be derived from the original letter |

#### Supplementary Table S4. Classification of physician corrections to LLM-generated patient-friendly letters

| Code | Category | Definition | n | % of corrections |
|---|---|---|---|---|
| 1 | Linguistic / stylistic | No change to the medical statement | 37 | 60.7 |
| 2 | Clarification | Content correct but too unspecific or ambiguous | 14 | 23.0 |
| 3 | Omission | Information missing | 4 | 6.6 |
| 4 | Factual inaccuracy | Incorrect medical statement | 5 | 8.2 |
| 5 | Misinterpretation | Original content transferred incorrectly | 0 | 0.0 |
| 6 | Unsupported addition | New statement without basis in the original letter | 1 | 1.6 |
| | **Total** | | **61** | **100.0** |

*Notes*. Safety relevance was assessed independently of correction type. None of the 61 documented corrections met the predefined criterion for a safety-relevant error. The additional omission identified after discharge was evaluated separately.

#### Supplementary Table S5. Patient characteristics of patients who completed T1 follow-up (n = 52)

| Characteristic | Group | Total n (%) |
|---|---|---|
| **Gender** | Female | 24 (46.2) |
| | Male | 28 (53.8) |
| **Age, years** | 18-35 | 7 (13.5) |
| | 36-45 | 4 (7.7) |
| | 46-55 | 4 (7.7) |
| | 56-65 | 10 (19.2) |
| | 66-75 | 15 (28.8) |
| | >75 | 12 (23.1) |
| | Median (IQR) | 66.5 (52.5-75.0) |
| **Disease** | Large vessel vasculitis | 18 (34.6) |
| | Rheumatoid arthritis | 10 (19.2) |
| | Connective tissue disease | 7 (13.5) |
| | ANCA-associated vasculitis | 5 (9.6) |
| | Polymyalgia rheumatica | 4 (7.7) |
| | Psoriatic arthritis | 2 (3.8) |
| | Still’s disease | 2 (3.8) |
| | Gout | 1 (1.9) |
| | IgG4-related disease | 1 (1.9) |
| | Reactive arthritis | 1 (1.9) |
| | Sarcoidosis | 1 (1.9) |
| **Education (n = 51)** | No education qualification | 1 (2.0) |

| | | |
|---|---|---|
| | Lower education qualification | 11 (21.6) |
| | Intermediate education qualification | 18 (35.3) |
| | Higher education qualification | 13 (25.5) |
| | University degree | 8 (15.7) |
| **Health literacy, eHEALS score** | Low | 11 (21.2) |
| | Moderate | 24 (46.2) |
| | High | 17 (32.7) |

#### Supplementary Table S6. Patient-reported information seeking about the discharge letter and physician contact before outpatient follow-up

| Survey item or response | Frequency |
|---|---|
| **Sought further information about the discharge letter** | **26/51 (51.0%)** |
| *Sources of further information* | |
| Internet | 24/51 (47.1%) |
| General practitioner | 14/51 (27.5%) |
| Friends and acquaintances | 10/51 (19.6%) |
| Other specialist physicians | 9/51 (17.6%) |
| Medical reference books | 5/51 (9.8%) |
| Other healthcare professionals | 5/51 (9.8%) |
| **Contacted a physician with questions before the outpatient follow-up visit** | **19/51 (37.3%)** |
| *Physicians contacted* | |
| General practitioner | 16/19 (84.2%) |
| Specialist physician | 2/19 (10.5%) |
| Ward physician | 2/19 (10.5%) |
| *Reasons for contacting a physician (free-text mentions)* | |
| Side effects | 4 |
| Blood test results | 3 |
| Medication | 3 |
| Corticosteroid tapering | 2 |
| Vaccination status | 2 |
| Advice on infections in the family | 1 |
| Appointment scheduling | 1 |
| Blood pressure | 1 |

| Survey item or response | Frequency |
|---|---|
| Bone density | 1 |
| Examination | 1 |
| Follow-up after stent placement | 1 |
| Questions (unspecified) | 1 |
| Referral form | 1 |
| Whom to contact about additional medication | 1 |

*Notes*. Values are n/N (%) for closed-ended items. Percentages for the two yes/no items and for information sources use all 51 respondents. Percentages for physician types use the 19 respondents who reported contact. Respondents could report multiple sources and physician types. Free-text reasons are shown as counts of mentions.

#### Supplementary Table S7. Exploratory associations with patient-reported evaluations: changes in text-based scores

##### Changes in text-based scores and patient-reported understandability and usefulness

| Text measure | Patient-reported outcome | n | ρ | 95% CI | p value | Holm-adjusted p |
|---|---|---|---|---|---|---|
| ΔPEMAT | Understandability | 51 | 0.09 | −0.19 to 0.35 | 0.522 | 1.000 |
| ΔPEMAT | Usefulness | 51 | −0.11 | −0.37 to 0.16 | 0.441 | 1.000 |
| ΔFRE | Understandability | 51 | −0.24 | −0.48 to 0.02 | 0.086 | 0.518 |
| ΔFRE | Usefulness | 51 | 0.10 | −0.15 to 0.35 | 0.483 | 1.000 |
| ΔWSTF | Understandability | 51 | 0.16 | −0.12 to 0.41 | 0.273 | 1.000 |
| ΔWSTF | Usefulness | 51 | 0.03 | −0.24 to 0.30 | 0.826 | 1.000 |

*Notes*. Spearman rank correlations are reported. Δ denotes the paired change in the respective score between original and patient-friendly letters. Outcomes were assessed at follow-up. Holm adjustment was applied across the six correlations. All analyses were exploratory; no correlation reached a Holm-adjusted p value <0.05.
*Abbreviations*: CI, confidence interval; FRE, Flesch Reading Ease (Amstad adaptation); PEMAT, Patient Education Materials Assessment Tool, understandability score; WSTF, first Vienna formula for non-fiction texts (Wiener Sachtextformel).

#### Supplementary Table S8. Exploratory associations with patient-reported evaluations: patient characteristics

##### Patient characteristics and patient-reported evaluations

| Patient characteristic | Patient-reported outcome | n | ρ | 95% CI | p value | Holm-adjusted p |
|---|---|---|---|---|---|---|
| Age | Understandability | 51 | −0.082 | −0.35 to 0.20 | 0.565 | 1.000 |
| Educational attainment | Understandability | 50 | 0.221 | −0.06 to 0.47 | 0.122 | 0.733 |
| eHEALS | Understandability | 51 | 0.397 | 0.13 to 0.61 | 0.004 | 0.035 |
| Age | Usefulness | 51 | 0.215 | −0.07 to 0.47 | 0.130 | 0.733 |

| Patient characteristic | Patient-reported outcome | n | ρ | 95% CI | p value | Holm-adjusted p |
|---|---|---|---|---|---|---|
| Educational attainment | Usefulness | 50 | −0.382 | −0.60 to −0.11 | 0.006 | 0.0498 |
| eHEALS | Usefulness | 51 | 0.101 | −0.18 to 0.37 | 0.479 | 1.000 |
| Age | Preference for future LLM letters | 51 | 0.154 | −0.13 to 0.41 | 0.279 | 1.000 |
| Educational attainment | Preference for future LLM letters | 50 | −0.247 | −0.49 to 0.04 | 0.084 | 0.590 |
| eHEALS | Preference for future LLM letters | 51 | −0.039 | −0.31 to 0.24 | 0.784 | 1.000 |

*Notes.* Spearman rank correlations are reported. Outcomes and eHEALS were assessed at follow-up. n indicates the number of observations included in each correlation. Holm adjustment was applied across the nine correlations. All analyses were exploratory; only the correlations between eHEALS and understandability and between educational attainment and usefulness reached a Holm-adjusted $p$ value <0.05.
*Abbreviations:* CI, confidence interval; eHEALS, eHealth Literacy Scale; LLM, large language model.
